# The Longevity-Frailty Hypothesis: Evidence from COVID-19 Death Rates in Europe

**DOI:** 10.1101/2020.04.14.20065540

**Authors:** Sammy Zahran, Levi Altringer, Ashok Prasad

## Abstract

By the end of spring (May 31st), the COVID-19 death rate was remarkably unevenly distributed across the countries Europe. While the risk of COVID-19 mortality is known to increase with age, age-specific COVID-19 death rates across Europe were similarly aberrantly distributed, implying that differences in age structure is an unlikely source of European variation in COVID-19 mortality. To explain these mortality distributions, we present a simple model where more favorable survival environments promote longevity and the accumulation of health frailty among the elderly while less favorable survival environments induce a mortality selection process that results in lower health frailty. Because the age-related conditions of frailty render the elderly less resistant to SARS-CoV-2, pre-existing survival environments may be non-obviously positively related to the COVID-19 death rate. To quantify the *survival environment* parameter of our model, we collected historic cohort- and period-based age-specific probabilities of death across Europe. We find strong positive relationships between survival indicators and COVID-19 death rates across Europe, a result that is robust to statistical control for the capacity of a healthcare system to treat and survive infected persons, the timing and stringency of non-pharmaceutical interventions, and the volume of inbound international travelers, among other factors. To address possible concerns over reporting heterogeneity across countries, we show that results are robust to the substitution of our response variable for a measure of cumulative excess mortality. Consistent with the intuition of our model, we also show a strong negative association between age-specific COVID-19 death rates and pre-existing all-cause age-specific mortality rates for a subset of European countries. Overall, results support the notion that variation in pre-existing frailty, resulting from heterogeneous survival environments, partially caused striking differences in COVID-19 death during the first wave of the pandemic.

## 1 Introduction

At the end of spring (May 31st), the COVID-19 death rate (deaths over population) was strikingly unevenly distributed across Europe (Figure 1). Among the hardest hit countries in the first wave of the pandemic were Belgium, Spain, Italy, and the United Kingdom with COVID-19 death rates of 816, 580, 553, and 552 deaths per 1 million, respectively.^1^ As of May 31st, 2020, the United Kingdom’s COVID-19 death rate exceeded the similarly developed economy of Germany (102 per million) by more than a factor of 5. Few epidemiological indicators distinguish the countries of Europe by such an extent.

**Figure 1:**
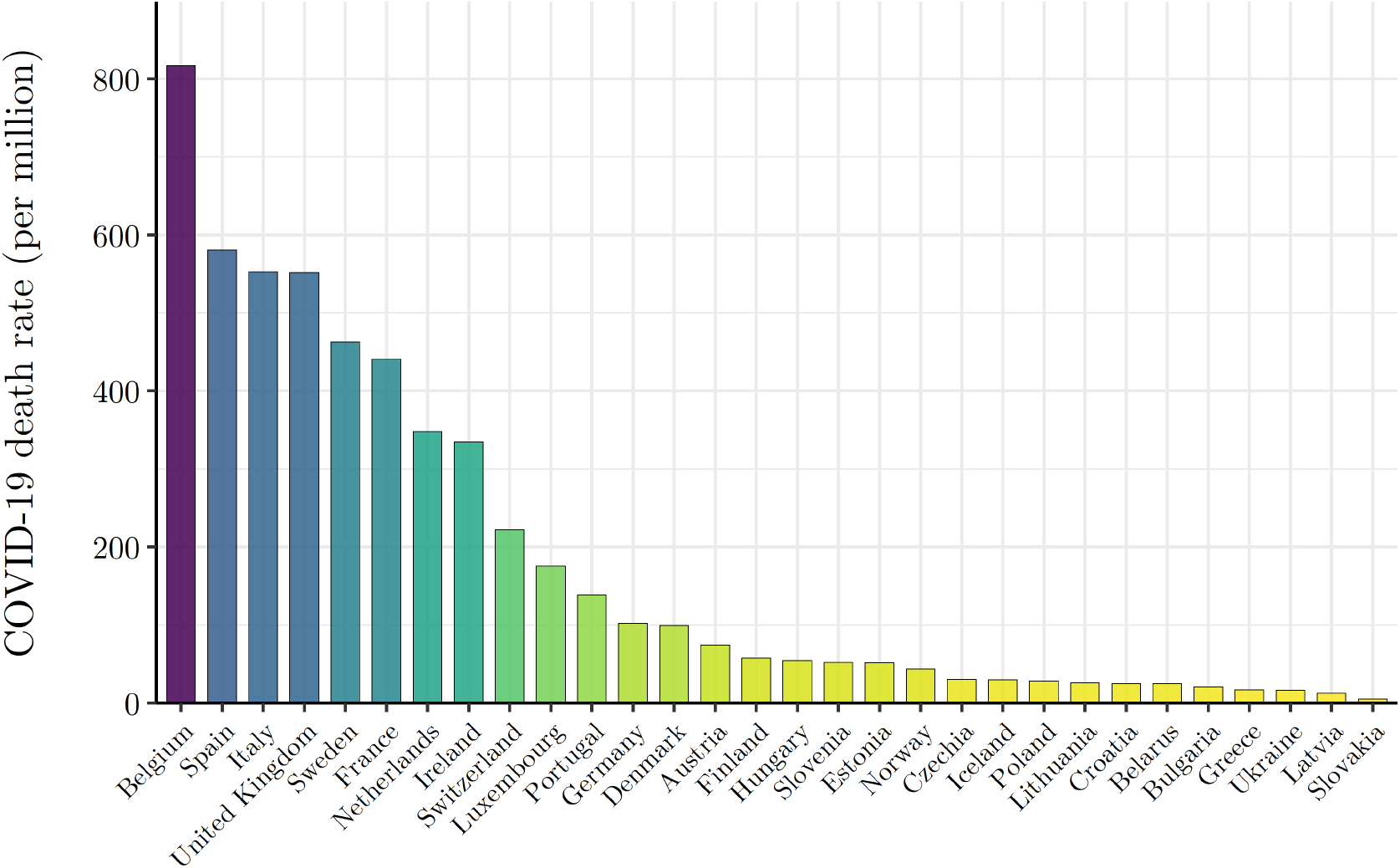
Uneven Distribution of Wave I (ending May 31st, 2020) COVID-19 Mortality Risk Across the Countries of Europe NOTE: The countries of Europe are arranged in descending order by reported COVID-19 deaths per million, as of May 31st, 2020. COVID-19 death data by country are from Johns Hopkins Coronavirus Resource Center (JHU CSSE) (https://github.com/CSSEGISandData/COVID-19_Unified-Dataset).

Because the risk of COVID-19 death is significantly higher among the elderly one might assume that variation in the COVID-19 death rate across Europe in this period reflects differences in age structure [2, 3]. Consider Figure 2 that plots age-specific COVID-19 death rates (as of May 31st, 2020) for nine European countries with readily available data. The size of each point plotted in Figure 2 reflects the share of total reported COVID-19 deaths that can be attributed to each age group within a country. Most striking is that the cross-national variation observed in Figure 1 remains. England’s age-specific death rates are roughly five times that of Germany’s across all age groups. The within-age-group variation in COVID-19 death rates across countries implies that the age structure of the population cannot account for observed differences in COVID-19 mortality risk between European countries. In fact, the percentage of the population ≥ 65 (*r* = 0.134, *p* = 0.480) and ≥ 75 (*r* = 0.186, *p* = 0.325) years of age are statistically uncorrelated with the COVID-19 death rate during this phase of the pandemic.

**Figure 2:**
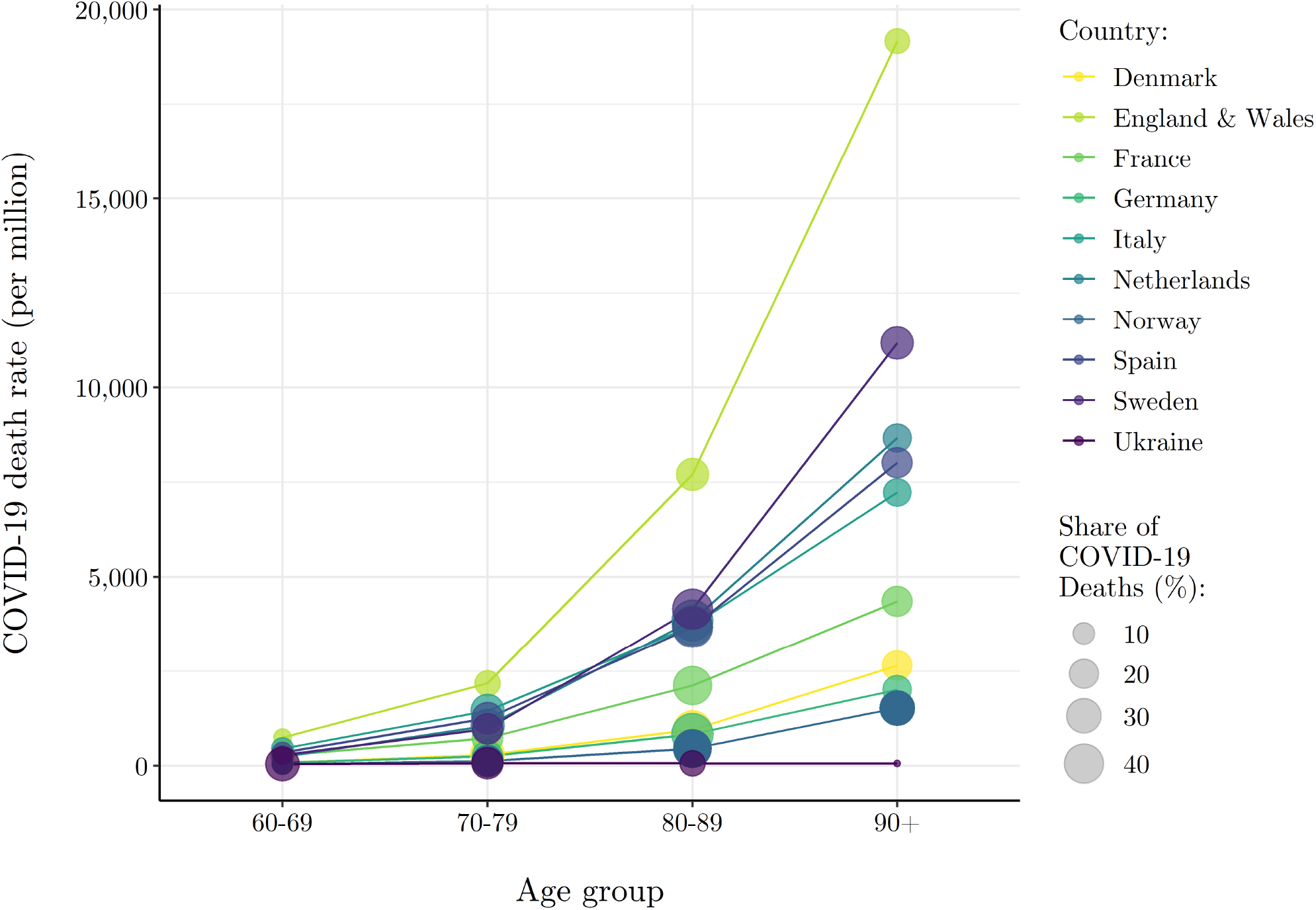
Heterogeneous Wave I (ending May 31st, 2020) COVID-19 Mortality Risk by Age Group Across Select European Countries NOTE: COVID-19 death and population data (closest to May 31st, 2020) by age and country are from National Institute for Demographic Studies (INED) (https://dc-covid.site.ined.fr/en/data/).

While percent elderly is statistically independent of the COVID-19 death rate, the risk of death by COVID-19 is manifestly related to age. Across the nine countries in Figure 2, persons 70 years of age and older account for more than 80 percent of all reported COVID-19 deaths. A theory of the puzzling distribution of the COVID-19 death rate across Europe at the end of spring must therefore account for the age-related nature of COVID-19 death risk.

Frailty is a technical notion used to describe a range of age-related conditions that render the elderly less resistant to health shocks [4, 5, 6, 7]. The accumulation of frailty in a society is non-obviously positively related to the survival environment [8, 9]. Societies characterized by higher pre-existing rates of elderly survival and consequent frailty accumulation may be more susceptible to higher rates of COVID-19 death, at least initially before the arrival of more effective treatment modalities.

Consider two similarly economically developed countries with varying rates of survival to age 75. Suppose that one country has high and the other a low survival environment with respect to all causes of death. Persons surviving to 75 in the low survival environment are more positively selected on the underlying ability to persist from one year to the next—i.e., longevity. Given positive mortality selection, elderly persons over the age of 75 in the low survival country may be more likely to withstand adverse health shocks than similarly aged persons in the high survival country because of lower underlying frailty. Similarly, persons surviving to 75 in a high survival environment are more negatively selected on the underlying ability to persist, rendering such persons less likely to withstand adverse health shocks than similarly aged persons in a low survival environment because of higher underlying frailty.

In this paper we develop a simple model of longevity and accumulated health frailty that captures the intuition above more formally. Each country begins with an identical underlying distribution of health frailty among the population. Then, heterogeneous survival environments across countries induce a mortality selection process that generates varying amounts of accumulated health frailty in subsequent periods. Specifically, more favorable survival environments promote longevity and a larger amount of accumulated health frailty while less favorable survival environments induce a relatively strict mortality selection process that results in a lower amount of accumulated health frailty among the elderly. With respect to an exogenous health shock that disproportionately targets the relatively health frail, like SARS-CoV-2, this simple model implies that countries with more favorable survival environments will have larger susceptible (health frail) populations and, therefore, higher COVID-19 death rates.

To measure a survival environment, we collected data on pre-existing all-cause age-specific death rates (measured in both period and cohort-specific terms), data of life expectancy at age 65, and develop an index of survival that is integrative of these measures. Across measures, we show that a more favorable pres-existing survival environment is strongly positively associated with the COVID-19 death rate across Europe. These results are robust to statistical control for competing theories of COVID-19 prevalence and mortality, including the capacity of a healthcare system to manage and survive infected persons, the timing and stringency of non-pharmaceutical interventions, and the volume of inbound international travelers. Results are also robust to the substitution of our response variable—the COVID-19 death rate— with a measure of cumulative excess mortality that addresses possible concerns of reporting heterogeneity across Europe.. For a subset of European countries with readily available data, and also compatible with our *longevity-frailty model*, we show a strong negative association between age-specific COVID-19 death rates and pre-existing all-cause age-specific mortality rates.

In the next section we describe our theoretical model outlining the relationship between a country’s survival environment and the expected accumulation of health frailty among the elderly. In Section 3 we detail data sources, variable operations and statistical models to test our model. In Section 4 we present results, including a series of robustness tests. In Section 5 we conclude with a summary of key findings and how our simple models extends to other facts of the COVID-19 pandemic.

## 2 Model

Consider a simple model of survival with two periods, *t* = (1, 2). In period *t* = 1, a country (*j*) begins with a representative population of individuals (*i*) who are each in possession of a level of health frailty (*f*_*i*_). Health frailty ranges from 0 (least frail) to 1 (most frail) and, for simplicity, is assumed to be distributed normally across the population with the restriction that *Pr*[0 ≤ *f* ≤ 1] = 1 since *f* ∈ [0, 1].

The likelihood that an individual survives to period *t* = 2 is determined by a Bernoulli trial. Specifically, individual *i* in country *j* survives to period *t* = 2 with probability *p*(*f*_*i*_; *δ*_*j*_) or, equivalently, dies in period *t* = 1 with probability 1 − *p*(*f*_*i*_; *δ*_*j*_). The survival probability function *p*(*f* ; *δ*), defined over the domain of health frailty, and dropping the subscripts for simplicity, takes the following functional form

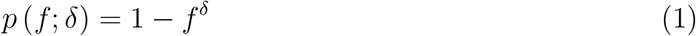

where *δ*> 1. Taking the first and second derivative of the survival probability function with respect to health frailty shows that the probability of survival increasingly deteriorates with health frailty

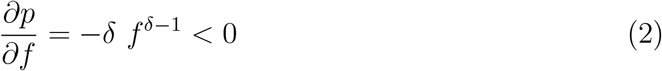

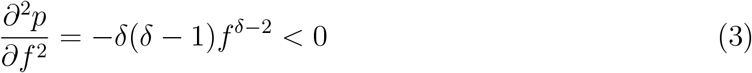

with certain survival for the least frail—*p* (0; *δ*) = 1—and certain death for most frail— *p* (1; *δ*) = 0. For a given level of health frailty in the range (0, 1) a more favorable survival environment increases an individual’s likelihood of surviving into older age. Specifically,

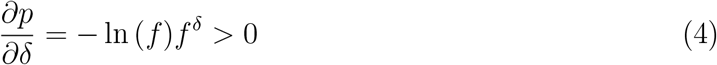

so that larger values of *δ* reflect a more favorable survival environment.

Given the survival probability function, we can derive the expected accumulation of health frailty in period *t* = 2. Let 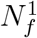 denote the number of individuals in period t=1 with health frailty f. Then the expected number of persons with health frailty *f* ≥ *h* that survive to period *t* = 2 is given by

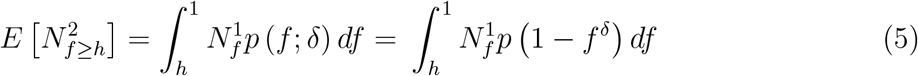

and the expected number of relatively frail individuals (*f* ≥ *h*) that survive to period t=2 is increasing in *δ*

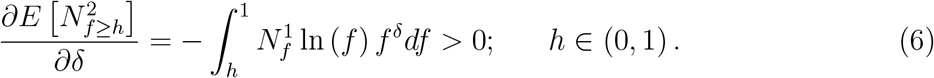

Thus, this simple model outlines a clear relationship between a country’s survival environment (*δ*) and the expected accumulation of health frailty among the elderly (period *t* = 2).^2^ Because frail persons are more susceptible to a range of age-related conditions that render them less resistant to health shocks or sudden changes to the survival environment like SARS-Cov2, we predict that higher survival environments are positively associated with higher COVID-19 death rates.

While we do not explicitly model the determination of *δ*, this environmental parameter is a function of several factors that contribute to the longevity of individuals across a health frailty distribution [13]. These factors may operate on populations in period-specific ways or cumulatively on a specific cohort over the life-course.^3^ In the next section, we develop and describe various period and cohort-based indicators of the survival environment used to test our model.

## 3 Data and Methods

### 3.1 Survival Environment Indicators

#### Cohort Survival

To obtain a cohort-based measure of survival environments across the countries of Europe we employ cohort death rate data from the Human Mortality Database (HMD).^4^ First, we subset the HMD data for countries in our sample to all cohorts born between 1910 and 1960—i.e., data for individuals at least 60 years of age in the year 2020— and collect *m*_*x*_, the observed death rate at age *x*. This generates different amounts of data for each country-cohort group. For instance, for the cohort born in 1920, data may be available for the age range of 0 to 106 years. For the cohort born in 1960, however, death rate data will only be available for the 0 to 56 years age range. Further, there is missing death rate data for some cohort-age observations for some countries. For the sake of comparability across countries, we subset the cohort *m*_*x*_ data to all cohort-age observations that are available for all countries in our sample.

To calculate *q*_*x*_ for each country-cohort-age group, which is defined as the probability that a person of age x will die within one year, we perform the following life table calculation

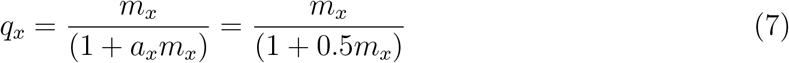

under the imposed assumption (shown above) that *a*_*x*_=0.5 for all age groups, which is consistent with the HMD Methods Protocols (pg. 36-37) [15]. Thus, for each country-cohort-age observation we are able obtain 1 − *q*_*x*_, which is the probability that a person of age *x* will survive to the following year. To get an average measure of the survival environment across cohorts for each country, we perform a LOESS regression of 1 − *q*_*x*_ on age to get 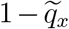 values.

To get an overall measure of the cohort-based survival environment across all age groups by country, we then perform the following calculation

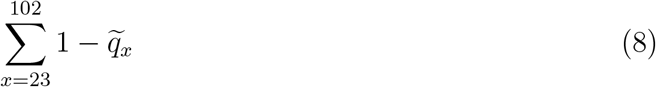

which is a discrete measure of the area under the 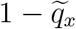 curve. For reference, across the 28 European countries in our sample for which cohort data are readily available—Germany and Croatia excluded—the minimum area calculated 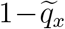 is 73.05 (Bulgaria), the maximum area calculated is 75.41 (France), and the mean and median areas are 74.62 and 74.37, respectively.

#### Period Survival

We employ the most recently available all-cause age-specific death rate data from HMD life tables—2016 for all countries except for Ukraine where the most recently available data are 2013—to obtain our first period-based measure of the survival environment for the countries of Europe. Specifically, for each country-age group—0 to 110 years for each European country—we collect *q*_*x*_, which, again, is defined as the probability that a person of age *x* will die within one year. Then, transforming this measure to 1 − *q*_*x*_ we obtain the probability that a person of age *x* will survive to the following year. Given that HMD measures terminate at 110 years of age, we impose the restriction that 1 − *q*_110_ = 0 for all countries.

Similar to our cohort-based measure of survival environment, we perform the area under the curve calculation

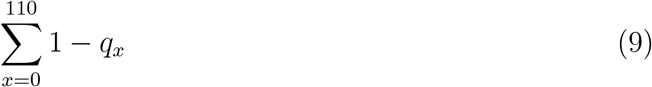

to get an overall measure of the survival environment across all age groups, by country. Across the 30 European countries in our sample, the minimum area calculated is 99.84 (Bulgaria), the maximum area calculated is 102.43 (France), and the mean and median areas are 101.42 and 101.55, respectively. In addition to the period-based measure presented above, we also employ the most recently available life table measure of *life expectancy at 65 years of age* as a period-based measure of survival environment. This is obtained directly through the HMD life tables.

#### Survival Index

Our three indicators of survival (described above) are all highly correlated. Thus, using Principal Component Analysis (PCA), we combine our cohort- and period-based measures of survival environment into one integrative index. Our results indicate that roughly 94 percent of the variation observed across these three measures is captured through the first principal component. The standardized predicted PCA score for each country in our sample serves as an additional indicator of survival to be employed in analyses of COVID-19 death risk across the countries of Europe.

#### Validation of Survival Environment Indicators

Our simple model outlined a clear relationship between a country’s survival environment and the accumulation of health frailty among the population. Insofar as our indicators of survival are an appropriate characterization of relative differences in survival environments across the countries of Europe, it follows that they can proxy, albeit roughly, for relative differences in the accumulation of health frailty.

Using data from the European Commission Eurostat database, we test the conceptual validity of our simple model and indicators of survival.^5^ Data are available for 14 of the 30 countries in our sample—see Appendix Table A1 for the list of countries where Germany is excluded given the absence of a cohort-based indicator of survival environment. We begin by constructing an elderly care index from three separate measures of elderly frailty; percent of disabled persons ≥ 65 years that need the assistance of others, percent of persons ≥ 65 years that need help with personal care activities, and percent of persons ≥ 65 years that need help with household activities. Employing PCA, we derive an integrative index of the need for elderly care using these measures. Across these three measures that proxy for elderly frailty and the need for care, the first principal component accounts for roughly 68 percent of overall variation.

In Appendix Table A1 we present scores for the three indicators of survival and the *Survival Index* alongside the three measures of elderly frailty and the *Elderly Care Index* for 13 similarly economically developed countries. The four countries that score highest according to our Survival Index — France, Spain, Italy, and the United Kingdom — also score highest on the Elderly Care Index, in the same order. Additionally, Iceland, Austria, and Finland rank in the bottom four countries across both indices. Further, in Appendix Figure A1, we present a correlation matrix which, using the data presented in Appendix Table A1, shows that each individual indicator of the survival environment and need for elderly care, as well as associated indices, are positively correlated with each other. We see this as an indication that our Survival Index, comprised of various indicators of the survival environment, proxy for relative differences in accumulated health frailty among the elderly across the countries of Europe.

### 3.2 COVID-19 Mortality Risk

Our measure of COVID-19 mortality risk comes from the John Hopkins University Center for System Sciences and Engineering COVID-19 Unified Dataset.^6^ Specifically, we combine COVID-19 death data with population data from the UN to calculate the daily cumulative COVID-19 death rate (deaths divided by population) over the period of March 4th, 2020 to May 31st, 2020, corresponding to the end of spring and the first wave of the pandemic.

We note here that the COVID-19 death rate may be biased by testing regimes and capabilities. The issue of testing bias is particularly problematic for our analysis if COVID-19 testing regimes and capabilities are correlated with survival environments. For instance, one might expect that countries with greater COVID-19 testing capabilities are likely to be the same countries that have the healthcare infrastructure necessary cultivate a relatively favorable survival environment. This would result in a positive relationship between the COVID-19 death rate and survival environment that is merely an illusion of testing capabilities. The standard econometric approach in this case would be to employ a fixed effects regression model, where unobserved heterogeneity in the response variable—such as testing capabilities—is controlled for through country fixed effects. However, given the time-invariant nature of our variables of interest, as well as the other control variables included in or model, we are unable to undertake this approach. Instead, we employ a second-best alternative—random effects regression model—which we discuss below. In the next section, we describe a series of control variables that account for these alternative explanations.

### 3.3 Control Variables

The control variables included in our empirical models account for alternative hypotheses of variation in COVID-19 mortality risk. These hypotheses include the timing and strength of epidemic seeding from exported cases, the capacity of healthcare systems to manage and survive infected persons, the stringency of non-pharmaceutical interventions (NPI), and whether or not a country requires mandatory Bacille Calmette-Guérin (BCG) vaccination.

To control for timing and strength of international seeding we collected the most recently available data from the World Bank on the annual count of inbound international tourists (2018). The intuition being that a country with a larger number of inbound international tourists would likely have a stronger initial seeding of the virus, which would likely influence the trajectory of the pandemic in each country. We also include World Bank data on hospital beds per capita (2015/2016), physicians per capita (2015/2016) and nurses per capita (2015/2016) to control for the capacity of healthcare systems in each country to manage and survive infected persons. Our measure of NPI stringency comes from the Oxford COVID-19 Government Response Tracker and is collected from the John Hopkins University Center for System Sciences and Engineering COVID-19 Unified Dataset. The Oxford COVID-19 Government Response Tracker collects information on several common policy responses that governments have implemented to respond to the pandemic, such as school closures, work-place closures, cancellation of public events, and stay at home orders to name a few. We use the Stringency Index which combines information about containment and closure policies with information about public information campaigns to generate a daily stringency score for each country that ranges from 0 to 100.

### 3.4 Statistical Models

Due to the time-invariant nature of our survival environment measures of interest, as well as the cross-sectional time-series nature of the data, we estimate versions of the following random effects model

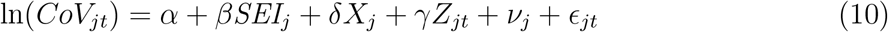

with

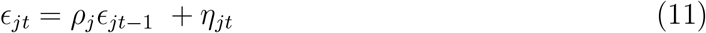

where *j* indexes country and *t* indexes day. *CoV* is the COVID-19 death rate. *SEI* is the survival environment indicator of interest, which, as we have previously shown, proxies for the accumulation, or build-up, of health frailty in a country. *X* is a vector of time invariant control variables, including the number of inbound international tourists, hospital beds per capita, physicians per capita, nurses per capita, and mandatory BCG vaccination status, while *Z* represents our only time-variant control, NPI policy stringency. *v*_*j*_ is a country-specific random effect and *E*_*jt*_ is the model disturbance term which is assumed to follow the AR(1) process shown above—where |*ρ*| < 1 and *η*_*jt*_ is independently and identically distributed (*i*.*i*.*d*.) with mean 0 and variance 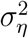.

We estimate three specifications of the general model presented above. First, we include our measures of interest, the indicators of survival environment (*SEI*), as a continuous variable — the natural log of life expectancy at 65 (ln *LE*_65_), the natural log of the cohort-based survival area (ln *CS*), the natural log of the period-based survival area (ln *PS*), and the natural log of the Survival Environment Index (ln *SI*)—so that

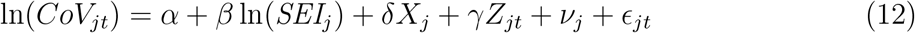

with *ε*_*jt*_ defined to follow the AR(1) process described by Eq. (11). According to our theoretical intuition that links favorable survival environments with larger amounts of accumulated health frailty, we expect the estimated *β* coefficient to be positive. In other words, we expect that countries with pre-existing survival environments that are relatively more favorable will have a higher COVID-19 death rate, all else equal. Specifically, the estimated *β* coefficient is interpreted as an elasticity. For example, in the case where the survival indicator is ln *LE*_65_, a 1 percent increase in life expectancy at 65 is associated with a *β* percent increase in the COVID-19 death rate, all else equal.

Second, we include the indicators of survival environment as a categorical variable by grouping our sample of European countries into terciles along the particular measure of interest so that

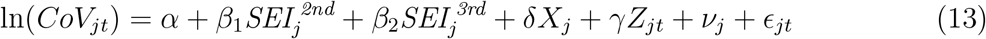

where 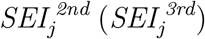 is a dummy variable equal to 1 if country *j* is in the second (third) tercile group for the survival environment indicator of interest—e.g., life expectancy at 65—and 0 otherwise. *ϵ*_*jt*_ follows the previously defined AR(1) process. Here, the reference group are all countries that fall into the first tercile range of the measure of interest. The interpretation of the *β* coefficients is less straight forward than in the previous specification. The exponentiated coefficient exp (*β*_1_) is the ratio of the expected geometric mean for the second tercile group over the expected geometric mean for the first tercile group, all else equal. Thus, [exp (*β*_1_) − 1] × 100 gives the associated percentage increase, or decrease if the difference is negative, in the COVID-19 death rate for the second tercile group relative to the first tercile group, all else constant [16].

Lastly, we continue with the categorical treatment of the Survival Index (*SI*) and interact it with week fixed effects to allow the estimated relationship between *CoV* and *SI* to vary over time. Specifically,

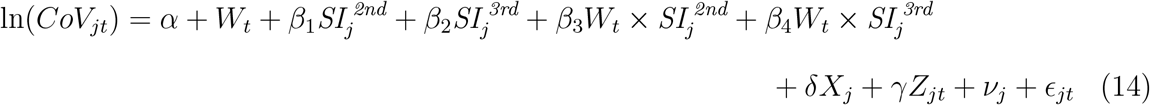

where *W*_*t*_ are the included week fixed-effects. In analyses that follow, we report the results of this model via post-estimation predicted margins, visually representing the divergence of COVID-19 deaths across survival environments in time.

## 4 Results

We begin with a simple assessment of the relationship between the COVID-19 death rate and our indicators of the survival environment. As expected by our model, we find strong positive correlations between the COVID-19 death rate and our indicators of the survival environment: life expectancy at 65 (*r* = 0.72, *p* < .001), period survival (*r* = 0.69, *p* < .001), cohort survival (*r* = 0.74, *p* < .001), and our survival index (*r* = 0.73, *p* < .001). In other words, higher pre-existing rates of elderly survival are associated with higher rates of COVID-19 death. Figure 3 displays the correlation spatially. Countries in yellow have the most unfavorable survival environments while countries in purple have the most favorable, according to our survival index. Hyper-imposed on the map are grey circles of varying size, corresponding to the observed COVID-19 death rate as of May 31st, 2020. The spatial correspondence between pre-existing elderly survival and the COVID-19 death rate is evident.

**Figure 3:**
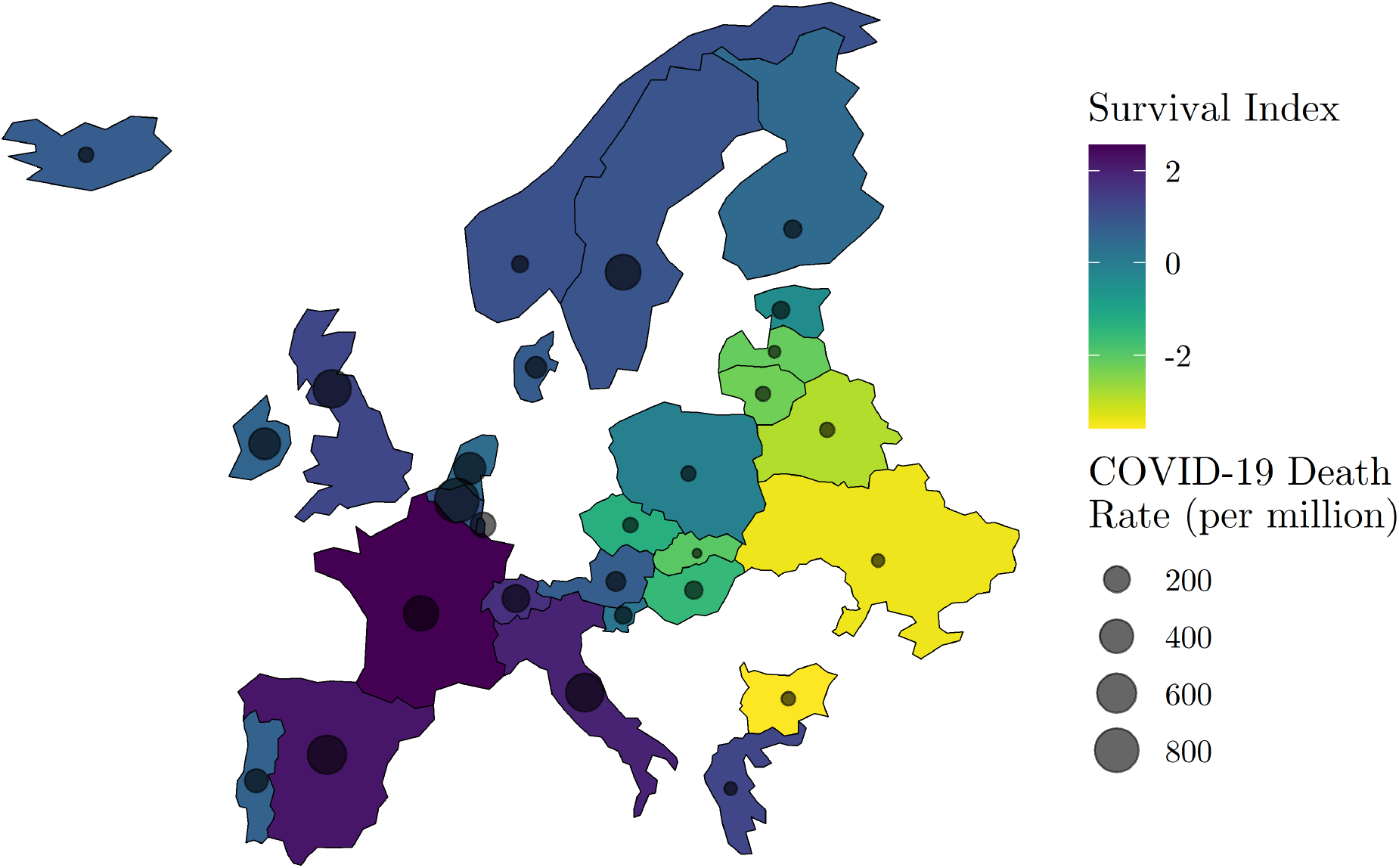
Spatial Distribution of Wave I (ending May 31st, 2020) COVID-19 Mortality Risk and Relative Survival Environment NOTE: COVID-19 death data by country are from Johns Hopkins Coronavirus Resource Center (JHU CSSE) (https://github.com/CSSEGISandData/COVID-19Unified-Dataset) and Survival Index data are derived from the Human Mortality Database (HMD) (https://www.mortality.org/).

Next, we test whether observed relationships between the COVID-19 death rate are robust to inclusion of variables that operationalize other candidate hypotheses, including timing and strength of epidemic seeding from exported cases, the capacity of healthcare systems to manage and survive infected persons, the stringency of non-pharmaceutical interventions (NPI), and whether or not a country requires mandatory Bacille Calmette-Guérin (BCG) vaccination. Importantly, our statistical models include a country random-effect and week fixed-effects to account for unobserved heterogeneity by place and time.

Table 1 reports regression coefficients from our random effects regression model (Eq.12) for all measures of the survival environment. The estimate of *β*, corresponding to our measures of the survival environment, is expected to be positive, indicating that higher pre-existing rates of survival among the elderly are associated with higher COVID-19 death rates. The necessary assumption for the identification of *β* is *E*[*v*_*j*_|SE_*j*_| = 0, with a similar condition being required for the identification of the coefficients on the other covariates. In other words, *v*_*j*_ is assumed to be the realization of an *i*.*i*.*d*. process with mean 0 and variance 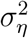.

As expected, all coefficients are positive and statistically distinguishable from chance. Focusing on column (4), corresponding to our integrative survival index (*SI*), we find that a standard deviation increase in the pre-existing survival environment increases the COVID-19 death rate by 52.2% (95% CI: 25.7, 78.8), other things held equal. A standard deviation increase in the survival index, for example, is the equivalent of moving from Greece (*SI* =1.28) to Spain (*SI* =2.22). The estimate of *ρ* is reported for all models in Table 1 along with the associated modified Bhargava et al. Durbin-Watson and Baltagi-Wu locally best invariant (LBI) test statistics under the null hypothesis *ρ* = 0 [17, 18]. This null hypothesis is rejected across all models presented in Table 1, and implying that the modeled AR(1) disturbance structure appropriately corrects the model estimated standard errors.

**Table 1:** Coefficients of Interest for Regression of *Ln* Cumulative COVID-19 Death Rate on *Ln* Survival Environment Indicators

|  | (1) | (2) | (3) | (4) |
| --- | --- | --- | --- | --- |
| Life Expectancy at 65 (LE) | 10.251***<br>(2.496) |  |  |  |
| Cohort Survival (CS) |  | 99.159***<br>(23.739) |  |  |
| Period Survival (PS) |  |  | 102.897***<br>(34.768) |  |
| Survival Index (SI) |  |  |  | 0.522***<br>(0.136) |
| Constant | -38.544***<br>(7.235) | -433.442***<br>(101.231) | -483.180***<br>(159.862) | -7.340***<br>(2.378) |
| $\hat{\rho}$ | 0.859 | 0.859 | 0.859 | 0.859 |
| Bhargava et al. Durbin-Watson | 0.382 | 0.375 | 0.382 | 0.375 |
| Baltagi-Wu LBI | 0.418 | 0.410 | 0.418 | 0.410 |
| Observations | 2,670 | 2,492 | 2,670 | 2,492 |
| Countries | 30 | 28 | 30 | 28 |
| Country RE | Yes | Yes | Yes | Yes |
| Week FE | Yes | Yes | Yes | Yes |
| Controls | Yes | Yes | Yes | Yes |

NOTE: Standard errors in parentheses. Levels of statistical significance are indicated by \*\*\* $p < 0.01$ , \*\* $p < 0.05$ , \* $p < 0.1$ . All models incorporate an AR(1) disturbance structure and the estimated $\rho$ is reported along with the associated modified Bhargava et al. Durbin-Watson and Baltagi-Wu locally best invariant (LBI) test statistics, both of which have complicated distributions [17, 18]. In all cases we reject the null hypothesis of $\rho = 0$ .

Figure 4 shows predicted COVID-19 death rates from Eq.(12) across percentile scores of all indicators of the pre-existing elderly survival environment. We derive predicted COVID-19 death rates by fixing other model covariates pertaining to inbound international travelers, healthcare system capacity, the stringency of non-pharmaceutical interventions (NPI), and week of observation are set at their sample means. We observe high statistical agreement across period and cohort-based operations of the elderly survival environment. Results indicate that the expected COVID-19 death rate is 3.7X higher for countries at the 75th percentile of our survival index as compared to countries at the 25th percentile. Overall, Figure 4 implies that the rate of COVID-19 mortality increased linearly in pre-existing elderly survival over the first wave of the COVID-19 pandemic.

**Figure 4:**
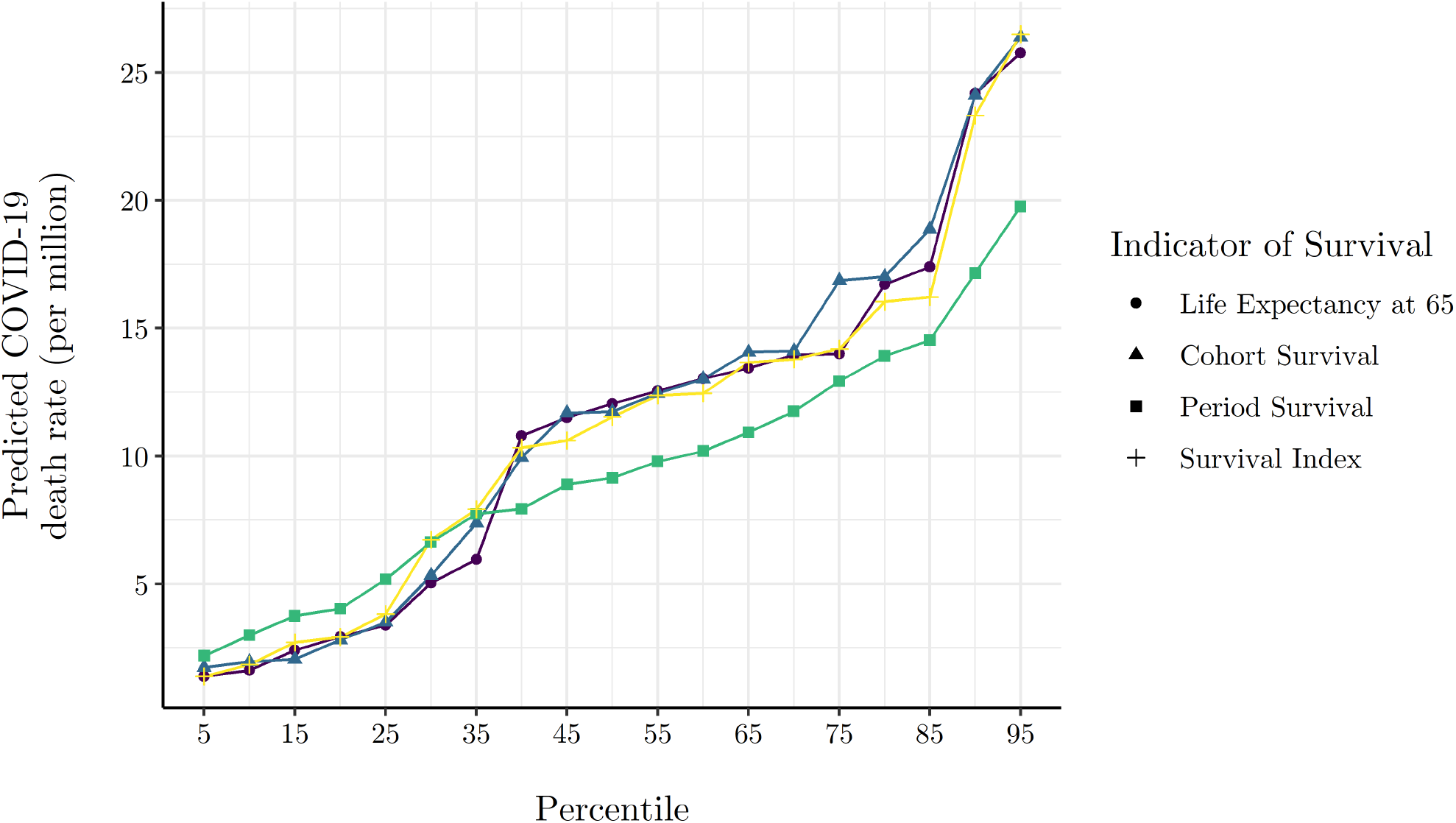
Model Predicted Cumulative COVID-19 Death Rate at Various Percentiles of Survival Environment Indicators. NOTE: Model predicted cumulative death rates are generated via the Eq. (12) model estimates presented in columns (1)-(4) of Table 1, where all other covariates are evaluated at their mean.

Table 2 reports coefficients from Eq.(13), where measures of the elderly survival environment are divided categorically into terciles of low, medium, and high survival. Because our response variable is measured in natural log terms, reported coefficients have the interpretation of a semi-elasticity reflecting the percent difference in the COVID-19 death rate of a tercile of interest relative to the reference tercile I of low pre-existing elderly survival. Consistent with theoretical expectation, coefficients across all operational definitions of the elderly survival environment are positive and statistically significant. Focusing on column (4), and all other things held equal, we find that countries in tercile II (medium survival environment) have an average COVID-19 death rate that is 444.7% higher (95% CI: 99.0, 1,379.1) than countries in tercile I (low survival environment).^7^ Similarly, and relative to tercile I (low survival environment), countries in tercile III (high survival environment) have an average COVID-19 death rate that is 784.6% higher (95% CI: 203.4, 2,473.9). Results in Table 2 corroborate Table 1 and Figure 4 implying that, other things held equal, the COVID-19 death rate increased monotonically with pre-existing elderly survival over the first wave of the pandemic.

**Table 2:** Coefficients of Interest for Regression of *Ln* Cumulative COVID-19 Death Rate on Categorical Survival Environment Indicators

|  | (1) | (2) | (3) | (4) |
| --- | --- | --- | --- | --- |
| LE Tercile II | 1.687***<br>(0.429) |  |  |  |
| LE Tercile III | 2.221***<br>(0.520) |  |  |  |
| CS Tercile II |  | 1.209***<br>(0.461) |  |  |
| CS Tercile III |  | 1.923***<br>(0.572) |  |  |
| PS Tercile II |  |  | 0.900**<br>(0.418) |  |
| PS Tercile III |  |  | 1.397***<br>(0.504) |  |
| SI Tercile II |  |  |  | 1.695***<br>(0.513) |
| SI Tercile III |  |  |  | 2.177***<br>(0.547) |
| Constant | -9.983***<br>(2.075) | -7.390***<br>(2.598) | -9.484***<br>(2.392) | -9.577***<br>(2.254) |
| $\hat{\rho}$ | 0.859 | 0.859 | 0.859 | 0.859 |
| Bhargava et al. Durbin-Watson | 0.382 | 0.375 | 0.382 | 0.375 |
| Baltagi-Wu LBI | 0.418 | 0.410 | 0.418 | 0.410 |
| Observations | 2,670 | 2,492 | 2,670 | 2,492 |
| Countries | 30 | 28 | 30 | 28 |
| Country RE | Yes | Yes | Yes | Yes |
| Week FE | Yes | Yes | Yes | Yes |
| Controls | Yes | Yes | Yes | Yes |

Next, Figure 5 presents results from Eq.(14), showing predicted cumulative COVID-19 death rates by Survival Index terciles over the course of the first wave of the European pandemic. Again, predictions are derived with all other model covariates fixed at their sample means. By mid-April, the predicted COVID-19 death rate in high elderly survival countries was almost 8X that observed in low elderly survival countries (69.4 vs 8.7 deaths per million). By the end of spring, the COVID-19 death rate of high elderly survival countries stood at 150 deaths per million, about 1.2X and 5.6X higher than medium and low survival environments, respectively.

**Figure 5:**
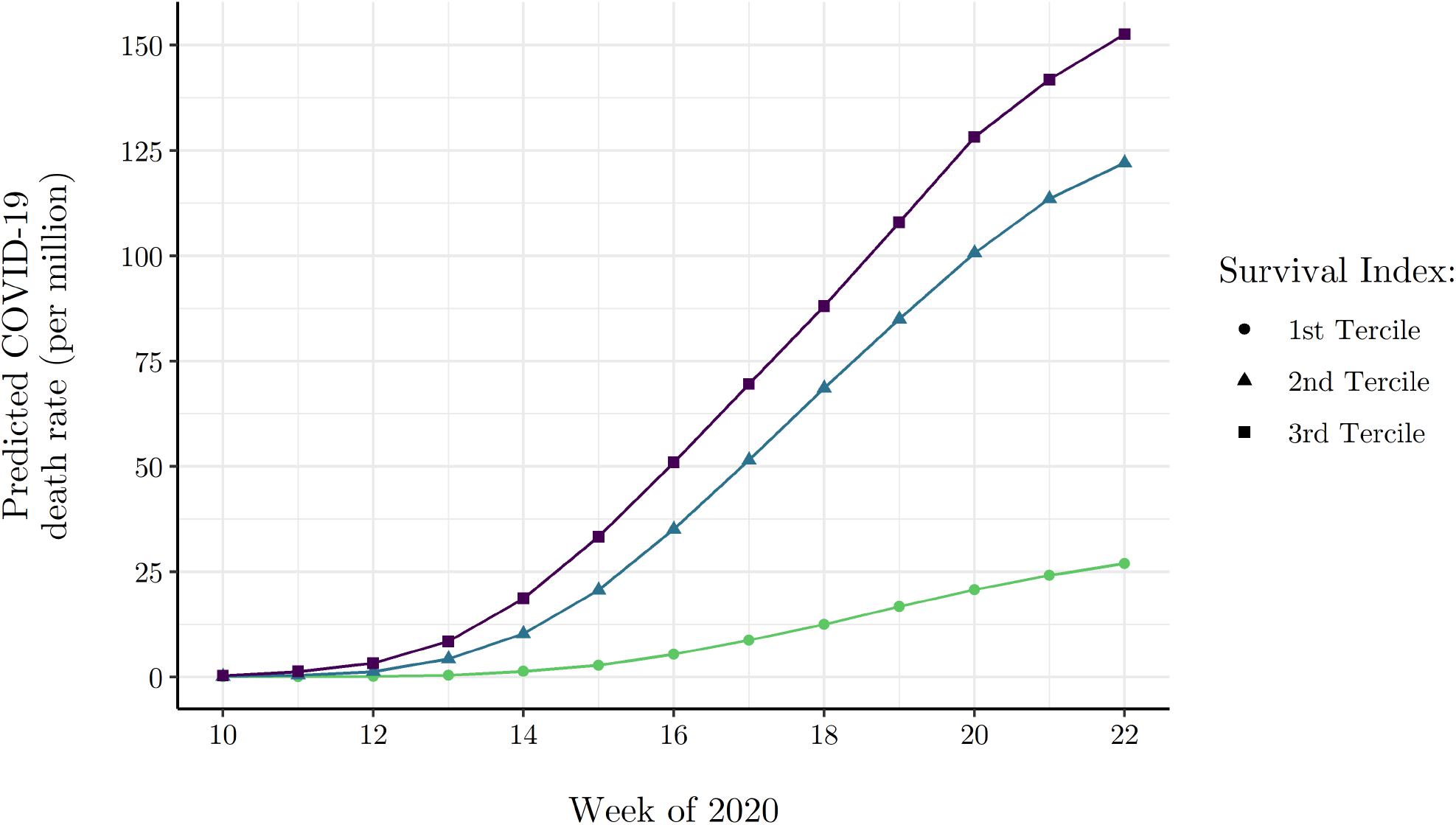
Model Predicted Cumulative COVID-19 Death Rate by Week of Wave I for Survival Index Terciles NOTE: Model predicted cumulative death rates by week and Survival Index tercile are generated via Eq. (14) model estimates, where all other covariates are evaluated at their mean.

## 5 Robustness

### 5.1 Excess Mortality

Given known inconsistencies in the reporting of COVID-19 deaths across countries—especially during the first wave of the pandemic—excess mortality can be used as a proxy for COVID-19 mortality that is potentially less biased by administrative differences that lead to under- or over-reporting of COVID-19 deaths. We obtained weekly excess mortality *Z*-scores from EuroMOMO ^8^. Each observation in the EuroMOMO data reflects the within country deviation from central tendency, or baseline, in all-cause mortality for a given week. Because of the within-country differenced nature of EuroMOMO’s excess mortality measure, we estimate following panel-corrected standard errors model

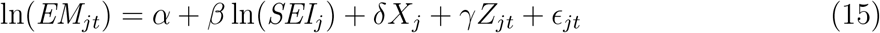

where all terms carry from Eq. (10), with the exception of our response variable *EM*_*jt*_ which measures the cumulative excess mortality of country *i* in week *j* in standardized terms. *E*_*jt*_ follows the same AR(1) disturbance structure defined in Eq. (11).

Table 3 reports the estimated coefficients. As before, our interpretive focus is on column (4) corresponding to our survival index. Because our survival index is measured in standardized terms, our coefficient of interest, *β*, has the interpretation of an elasticity, indicating that a standard deviation increase in the pre-existing survival environment is associated with a *β* percent change in excess mortality. Other things held equal, we find that a unit increase in our survival index is associated 6.1% increase (95% CI: 1.5, 10.7) in cumulative weekly excess mortality.

**Table 3:** Coefficients of Interest for Regression of *Ln* Cumulative Excess Mortality on *Ln* Survival Environment Indicators

|  | (1) | (2) | (3) | (4) |
| --- | --- | --- | --- | --- |
| Life Expectancy at 65 (LE) | 3.202***<br>(0.925) |  |  |  |
| Cohort Survival (CS) |  | 32.305***<br>(10.460) |  |  |
| Period Survival (PS) |  |  | 31.048***<br>(11.311) |  |
| Survival Index (SI) |  |  |  | 0.061***<br>(0.023) |
| Constant | -10.219***<br>(3.599) | -139.669***<br>(45.759) | -143.971***<br>(53.065) | -0.953<br>(0.985) |
| $\hat{\rho}$ | 0.821 | 0.833 | 0.836 | 0.841 |
| Observations | 247 | 234 | 247 | 234 |
| R-squared | 0.701 | 0.725 | 0.703 | 0.724 |
| Countries | 19 | 18 | 19 | 18 |
| Week FE | Yes | Yes | Yes | Yes |
| Controls | Yes | Yes | Yes | Yes |

NOTE: Standard errors in parentheses. Levels of statistical significance are indicated by \*\*\* $p < 0.01$ , \*\* $p < 0.05$ , \* $p < 0.1$ . All models incorporate an AR(1) disturbance structure and the estimated $\rho$ is reported.

### 5.2 Age-Specific Mortality

The National Institute for Demographic Studies (INED) collects age- and sex-specific COVID-19 death data for select European countries.^9^ We collapse age-specific COVID-19 deaths into common age intervals across the countries for which data are available and calculate corresponding pre-existing all-cause mortality rates using data from the HMD. Age-specific COVID-19 death data is available for all age groups in England, France, Germany, Italy, the Netherlands, Spain, Sweden and Ukraine. Denmark’s age-specific data is only available for those 60 years of age and older, Norway data is available for those 40 years of age and older, and Portugal’s data is top-coded at 80 years of age so that comparable data is only available for those 79 years of age and younger. Pre-existing age-specific all-cause mortality data are anchored on the most recently available common year of 2016. To test the statistical relationship between pre-existing all-cause mortality and the COVID-19 death rate, we estimate the following least squares model

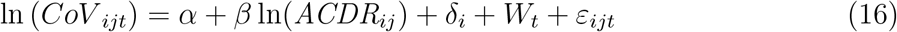

where *CoV* is the COVID-19 death rate for age group *i* in country *j* at time *t, ACDR* is the pre-existing all-cause death rate (2016), *δ*_*i*_ is a vector of age-interval fixed effect, and *W*_*t*_ is a vector of week fixed effects. *ε* is the model disturbance term. Our theoretical interest is in the estimate of *β* which we expect to be negative, implying that, other things equal, a 1 percent increase in the age-specific all-cause death rate is associated with a *β* percent decrease in the associated age-specific COVID-19 death rate.

Table 4 reports the coefficient estimates for equation (15). The model estimates presented in column (1) are generated in the absence of age-interval fixed effects. In columns (2) through (4) we introduce age-interval fixed effects. The inclusion of these variables results in a between-country estimate of that is purged of the confounding within-country effect of increasing risk of death across age intervals. According to our theoretical expectations, pre-existing all-cause mortality rates are estimated to have a strong, negative association with the COVID-19 death rate across countries. The coefficient estimate of −1.439 (95% CI: −1.519, −1.360) in column (2) implies that, other things equal, a 1% increase in the pre-existing all-cause death rate is associated with an approximate 1.5 percent lower COVID-19 death rate, on average across age intervals.

In column (3) of the table we reduce the sample to intervals ≥ 60 years of age. The qualitative nature of the estimates are maintained, though the negative association between pre-existing all-cause mortality and the COVID-19 death rate appears to be stronger among the elderly. Specifically, a 1% increase in the pre-existing all-cause death rate is associated with an approximate 4.3 percent decrease in the COVID-19 death rate for those ≥ 60 years of age, other things equal. In column (4) we restrict the sample to intervals ≤ 49 years of age. Relative to column (3), which restricts to intervals ≥ 60 years of age, the coefficient estimate in column (4) intuitively deflates, reinforcing that the pre-existing death rate effect is stronger among the elderly and compatible with a longevity-frailty model.

**Table 4:** Regression of *Ln* Cumulative Age-Specific COVID-19 Death Rates on *Ln* Pre-Existing All-Cause Age-Specific Death Rates

|  | (1) | (2) | (3) | (4) |
| --- | --- | --- | --- | --- |
| All-Cause Death Rate (2016) | 1.112***<br>(0.010) | -1.439***<br>(0.041) | -4.313***<br>(0.095) | -0.653***<br>(0.034) |
| Age 10-19 |  | -0.758***<br>(0.104) |  | -0.169**<br>(0.072) |
| Age 20-29 |  | 1.664***<br>(0.089) |  | 1.600***<br>(0.061) |
| Age 30-39 |  | 3.595***<br>(0.093) |  | 3.001***<br>(0.065) |
| Age 40-49 |  | 5.570***<br>(0.106) |  | 4.364***<br>(0.078) |
| Age 50-59 |  | 8.101***<br>(0.132) |  |  |
| Age 60-69 |  | 10.408***<br>(0.160) |  |  |
| Age 70-79 |  | 12.831***<br>(0.192) | 5.009***<br>(0.104) |  |
| Age 80-89 |  | 15.571***<br>(0.234) | 10.979***<br>(0.201) |  |
| Age $\geq 90$ | | 17.749***<br>(0.274) | 16.161***<br>(0.297) | |
| Constant | 4.843***<br>(0.501) | -17.480***<br>(0.487) | -20.587***<br>(0.649) | -10.379***<br>(0.581) |
| Observations | 4,697 | 4,697 | 2,335 | 1,820 |
| R-squared | 0.738 | 0.869 | 0.710 | 0.764 |
| Week FE | Yes | Yes | Yes | Yes |
| Age Interval FE | No | Yes | Yes | Yes |

NOTE: Standard errors in parentheses. Levels of statistical significance are indicated by \*\*\* $p < 0.01$ , \*\* $p < 0.05$ , \* $p < 0.1$ . For columns (2) and (4) the reference age group is 0-9. For column (3) the reference age group is 60-69.

## 6 Conclusion

In this paper we developed a simple model of longevity and accumulated health frailty in an attempt to explain observed differences in COVID-19 mortality across the countries of Europe. The model describes how heterogeneous survival environments across countries induce mortality selection processes that generate varying amounts of accumulated health frailty among elderly populations. With respect to an exogenous health shock that disproportionately targets the relatively health frail, like SARS-CoV-2, our model implies that countries with a more favorable survival environment will have a larger susceptible (health frail) population and, therefore, a higher COVID-19 death rate. To operationalize a country’s survival environment we collected data on pre-existing all-cause age-specific death rates (measured in both period and cohort-specific terms), life expectancy at age 65, and an index of survival that is integrative of these variables. We showed that our various indicators of the survival environment are correlated with measures of elderly frailty, serving as sound proxies of relative differences in accumulated health frailty among the countries of Europe.

We then statistically linked cross-national variation in survival environments to the puzzling distribution of COVID-19 death rates in Europe at the end of spring, finding that a more favorable survival environment, as indicated by lower pre-existing all-cause mortality rates, is positively associated with COVID-19 death rates. Specifically, we find that a standard deviation increase in the pre-existing survival environment increases the COVID-19 death rate by 52.2% (95% CI: 25.7, 78.8), other things held equal. Coupled with analyses involving categorical measurement of the elderly survival environment, the COVID-19 death rate appeared to increase monotonically with pre-existing elderly survival over the first wave of the pandemic. Importantly, these relationships are robust to statistical control for the inbound international travelers, the capacity of a healthcare system to manage and survive infected persons, and the timing and stringency of non-pharmaceutical interventions, among other factors.

We followed with analyses of cumulative excess mortality instead of the COVID-19 death rate. Results behaved similarly, implying that reporting differences across European countries is an unlikely source of our observed correlations between pre-existing survival environments and COVID-19 mortality. We closed analyses showing a strong negative association between age-specific COVID-19 death rates and pre-existing all-cause age-specific mortality rates for a subset of European countries with readily available data.

Our results are consistent with other known facts concerning COVID-19 mortality during the first wave of the pandemic. Specifically, end-of-spring data show that a large percentage of COVID-19 deaths occurred among frail populations in long-term care facilities — Belgium (53%), Denmark (34%), France (51%), Germany (36%), Hungary (19%), Ireland (60%), Norway (60%), Portugal (40%), and Sweden (45%).^10^ In France, Spain and Italy, among the hardest hit countries of Europe, about half to two-thirds of elderly require assistance for basic personal and household care activities. This fact educates that a sizeable percentage of European elderly could not easily socially distance in the first wave of the pandemic, compounding the risk of COVID-19 mortality.

Further, there are several characteristics of COVID-19 that explain why mortality could be strongly associated with frailty, as we have shown here. Firstly, persons with comorbidities such as type II diabetes, hypertension, cardiovascular disease, chronic kidney disease etc., especially those with multiple comorbidities, face a higher risk of death. Age > 80 years is also independently associated with high mortality [20, 21, 22, 23]. Secondly, while we now know that airborne transmission is the main mode of the spread of this virus [24, 25], this was not appreciated in the early months of the pandemic and public health advice on airborne transmission was inadequate, if not misleading [26, 27]. Thirdly, evidence suggests that pre-symptomatic and asymptomatic infected persons can and did spread the infection [28, 29]. Finally, the dynamics of viral shedding by infected people allow them to easily infect multiple people in closed or crowded environments and super-spreader events played an important role in early transmission dynamics [30, 31]. These factors proximately underwrite the association between mortality and the accumulation of frailty reported here.

In sum, our results support the hypothesis that variation in pre-existing frailty, stemming from differences in pre-existing survival environments, partially determined the striking differences in COVID-19 death risk during observed during the first wave of the European pandemic. Our findings also provide general intuition as to how similarly novel causes of death might operate across countries via the heterogeneous accumulation of health frailty among their populations. It is interesting that some reports suggest that the aged population in developing countries show a lower COVID-19 mortality rate than seen in Europe, which would be predicted by our model due to the poor survival environment that the aged face in these countries [32].

## Data Availability

Data and codes are available freely from the authors on request. All data used for the analysis is publicly available and links to data sources are provided in the manuscript.

## Appendix

**Table A1:** Survival and Elderly Care indexes by Country

| Country | Survival Index | Cohort Survival | Period Survival | Life Expectancy at 65 | Elderly Care Index | % > 65 Disabled Need Help | % > 65 Need Help Personal Activities | % > 65 Need Help House Activities |
| --- | --- | --- | --- | --- | --- | --- | --- | --- |
| France | 2.557 | 1.522 | 1.590 | 1.404 | 2.270 | 1.400 | 0.678 | 1.839 |
| Spain | 2.220 | 1.271 | 1.410 | 1.249 | 2.258 | 1.496 | 1.266 | 1.149 |
| Italy | 1.975 | 1.172 | 1.208 | 1.125 | 2.208 | 1.816 | 0.810 | 1.185 |
| United Kingdom | 1.259 | 0.778 | 1.004 | 0.476 | 1.115 | 0.746 | 0.611 | 0.575 |
| Norway | 1.040 | 0.519 | 0.558 | 0.803 | -1.486 | -0.890 | -1.076 | -0.613 |
| Luxembourg | 0.978 | 0.192 | 0.744 | 0.839 | -1.144 | -0.362 | -1.217 | -0.418 |
| Belgium | 0.969 | 0.574 | 0.608 | 0.571 | 1.394 | 0.623 | 0.310 | 1.470 |
| Sweden | 0.951 | 0.578 | 0.402 | 0.744 | -0.415 | -0.528 | -0.219 | 0.029 |
| Denmark | 0.805 | 0.370 | 0.843 | 0.256 | -0.449 | -0.785 | 0.464 | -0.437 |
| Iceland | 0.776 | 0.970 | -0.015 | 0.458 | -1.525 | -0.944 | -0.905 | -0.794 |
| Austria | 0.762 | 0.489 | 0.323 | 0.583 | -1.514 | -0.814 | -0.840 | -0.967 |
| Finland | 0.468 | 0.372 | -0.024 | 0.536 | -2.158 | -1.460 | -0.949 | -1.322 |
| Netherlands | 0.416 | 0.440 | -0.071 | 0.423 | -1.114 | -0.873 | -0.822 | -0.239 |

NOTE: This table presents standardized scores for three indicators of survival environment and the “Survival Index” alongside three measures of elderly frailty and the “Elderly Care Index” for 13 similarly economically developed countries. Survival environment indicator data are from the Human Mortality Database (HMD) (https://www.mortality.org/) and elderly frailty indicator data are from European Commission Eurostat database (https://ec.europa.eu/eurostat/data/database).

**Figure A1:**
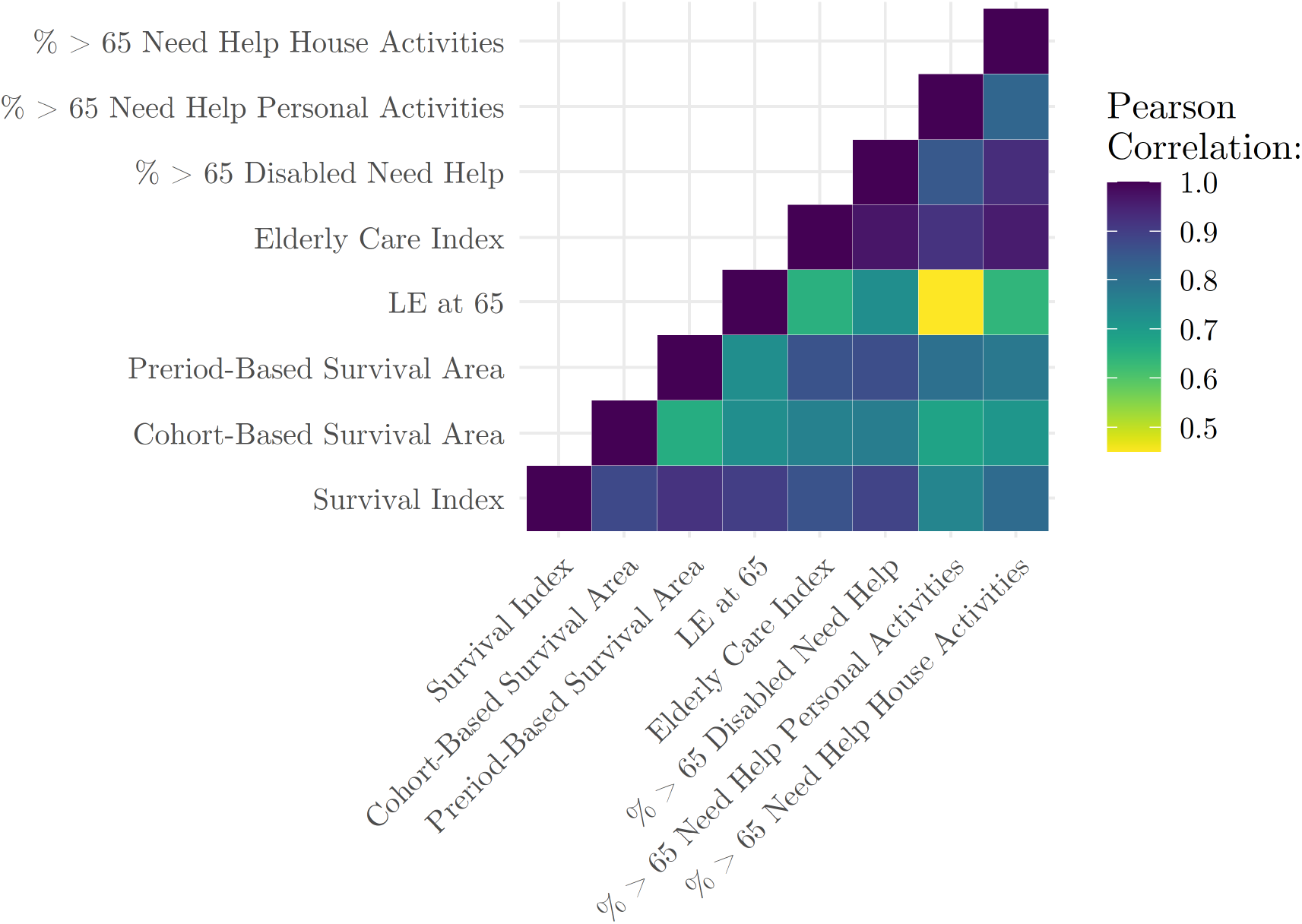
Correlation Matrix for Indicators of Survival Environment and Elderly Frailty NOTE: Survival environment indicator data are derived from the Human Mortality Database (HMD) (https://www.mortality.org/) and elderly frailty indicator data are from European Commission Eurostat database (https://ec.europa.eu/eurostat/data/database). The data underlying the correlations presented here can be found in Appendix Table A1.

## Footnotes

1 Reporting suggests that Belgium’s method of recording COVID-19 deaths includes not only deaths that are confirmed to be virus-related, but also deaths that are suspected of being linked, whether the victim was tested or not [1].

2 It is important to note that an individual’s health frailty is not endogenously determined in this simple model. For instance, a less favorable survival environment may positively contribute to the accumulation of health frailty through “scarring” or “weathering” effects even though the process of mortality selection negatively contributes to accumulated health frailty, or what some refer to as the “culling” effect [10]. A model that incorporates these competing effects would provide a more general theoretical understanding of accumulated health frailty that could account for within-country variation in health outcomes and/or mortality across groups—for example, the relatively high COVID-19 death rates observed for African Americans in the US [11] or the positive association between COVID-19 death risk and multiple deprivation in the UK [12].

3 For example, the period-based influenza hypothesis argues that COVID-19 death rates are higher Sweden than neighboring Nordic countries because Sweden experienced remarkably lower death rates from flu over the last seasons [14]. The period-specific attenuation of influenza mortality produced relatively higher rates of frailty that rendered the elderly in Sweden more susceptible to COVID-19 death.

4 HMD data were accessed at https://www.mortality.org/.

5 Eurostat data were accessed at https://ec.europa.eu/eurostat/data/database.

6 JHU CSSE data were accessed at https://github.com/CSSEGISandData/COVID-19_Unified-Dataset.

7 To obtain the marginal effects, in percentage terms, for the estimated coefficients in Table 2, one must perform the following transformation: [exp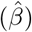 − 1] × 100 [16].

8 Excess mortality *Z*-scores were accessed at https://www.euromomo.eu/graphs-and-maps.

9 INED age- and sex-specific COVID-19 death data were accessed at https://dc-covid.site.ined.fr/en/data/.

10 These data come from the International Long-Term Care Policy Network [19] (https://ltccovid.org/wp-content/uploads/2020/05/Mortality-associated-with-COVID-3-May-final-5.pdf) and Sciensano, a public research institution which publishes very detailed epidemiological daily reports on COVID-19 (https://covid-19.sciensano.be/sites/default/files/Covid19/covid-19_daily_report20200505_-_fr.pdf).

